# Systematic Bias in Clinical Decision Instrument Development: A Quantitative Meta-Analysis

**DOI:** 10.1101/2025.02.12.25320965

**Authors:** Jed Keenan Obra, Chandan Singh, Kenshata Watkins, Jean Feng, Ziad Obermeyer, Aaron Kornblith

**Affiliations:** University of California, Berkeley; University of California, San Francisco; Microsoft Research

**Keywords:** Clinical decision rule, Race, Ethnicity, Gender, Sex, Bias

## Abstract

Clinical decision instruments (CDIs) face an equity dilemma. On the one hand, they often reduce disparities in patient care through data-driven standardization of best practices. On the other hand, this standardization may itself inadvertently perpetuate bias and inequality within healthcare systems. Here, we quantify different measures of potential for implicit bias present in CDI development that can inform future CDI development. We find evidence for systematic bias in the development of 690 CDIs that underwent validation through various analyses: self-reported participant demographics are skewed—e.g. 73% of participants are White, 55% are male; investigator teams are geographically skewed—e.g. 52% in North America, 31% in Europe; CDIs use predictor variables that may be prone to bias—e.g. 13 CDIs explicitly use *Race and Ethnicity*; outcome definitions may further introduce bias—e.g. 28% of CDIs involve follow-up, which may disproportionately skew outcome representation based on socioeconomic status. As CDIs become increasingly prominent in medicine, we recommend that these factors are considered during development and clearly conveyed to clinicians using CDIs.

## Introduction

Clinical decision instruments (CDIs) are tools used in healthcare to assist clinicians in diagnosing conditions, predicting patient outcomes, or guiding treatment decisions based on clinical data. These instruments are widely used with the aim of improving patient care through data-driven standardization.^1,2^ While CDIs generally enhance clinical care, this standardization may inadvertently perpetuate inequality within healthcare systems.^3–6^ As data-driven CDIs become more prominent in medicine, care must be taken to ensure that they not perpetuate (implicit) ***bias***—meaning that these tools should not unintentionally provide less accurate or effective guidance for patients across different socio-economic, racial, ethnic, or other demographic groups, as shaped by systemic inequities and social constructs.

Various factors can introduce implicit bias during the CDI development process. Perhaps most importantly, failing to enroll diverse populations in development cohorts can lead to inaccuracies, disproportionately affecting minority groups.^7^ Beyond enrollment, bias can arise from the predictor variables included in the CDI (e.g. *Race*)^7–9^ or in the definition of the outcome itself, which may skew the representation of different groups.^10,11^ Similarly, the regional affiliations and demographics of investigator teams may influence the perspectives and biases embedded in these instruments.^12^

Here, we take a closer look at these disparate factors contributing to CDI bias. Specifically, we analyze the CDIs in MDCalc^13^, an online tool that hosts hundreds of widely used CDIs. We find potential for bias introduced through four factors: patient cohort demographics, predictor variables, outcome definitions, and author demographics. Going forward, we recommend that these factors (in addition to standard best practices^14^) are conveyed clearly to clinicians, e.g. in a model card.^15^ Moreover, the CDI development process for new CDIs should carefully consider these sources of implicit bias in order to design equitable CDIs that work for diverse populations.

## Methods

### Inclusion criteria for primary studies on CDIs

To identify signals of bias within CDI development, we analyzed studies detailing the procedures by which CDI developers used to create their instruments. Using web data extraction, we collected studies from the MDCalc website (MDCalc.com, July 26, 2023) under the *Evidence* and *Original/Primary Reference* sections. We then manually searched and retrieved CDI primary literature using PubMed with the same title, authorship, and Digital Object Identifier (DOI). For studies that were not available on PubMed, we used the Bioscience, Natural Resources & Public Health Library of University of California, Berkeley to find full-texts of the primary references. Two team members reviewed each text to confirm primary literature of CDI. We excluded primary development literature that (i) was not written in English, (ii) exclusively described a validation study, (iii) used a textbook reference or established guideline (DSM-V) was used instead of a development research study, or (iv) was not available in library databases. Methodological standards were verified with the Prisma 2020 statement paper for systematic reviews of studies^16^.

### Characterizing participant demographics in primary studies

To identify subject characteristics that may be overrepresented, we collected participant count, biological sex, and racial/ethnic demographic info of participant cohorts. Keywords include, *Asian, Caucasian, Hispanic, Latino, African*, etc. To verify the accuracy of web extraction data and missing information, we verified participant demographic data by direct examination of the article. The keywords *male, female, sex*, and *gender* were manually searched to identify count and biological sex. Primary literature uses both *sex* and *gender* terminology to describe biological sex of their participant cohorts as well as *race* and/or *ethnicity* to describe groups of participants connected by common descent. Aligned with the Journal of the American Medical Association (JAMA)^17,18^, we solely use the term *sex* to report biological factors, *gender* to report gender identity, and *race and ethnicity* to describe major groupings assigned in the literature while recognizing the origin of these terms as social constructs differentiating common descent and cultural identity.

### Characterizing investigator demographics in primary studies

Authors, affiliations, and year of publication for each primary reference CDI study were identified from Pubmed. We excluded 125 CDIs in which the author’s name could not be analyzed (e.g. only initials available on authorship creditation). We used ChatGPT (GPT-3.5-Turbo, accessed through OpenAI’s python interface^19^ to automatically infer the gender identity of each author and to identify the country name corresponding to their affiliation. Specifically, we prompt ChatGPT with the input, *Return whether the name ‘{name}’ is more common for a male or a female. Answer with one word, ‘Male’ or ‘Female’*. To ensure that ChatGPT sex categorizations are reliable, the ChatGPT output was validated by matching the gender pronouns given on an author’s homepage. We searched 150 randomly selected author name/affiliation pairs (75 predicted as male, and 75 predicted as female) and 81 ground truth author sex were available from author homepages. 80 of 81 (99%) author sexes were correctly identified by ChatGPT.

### Predictor variables and categorization

We used curated information from MDCalc contributors to identify CDI predictor variables and classify CDI by disease, system, purpose, chief complaint, and specialty. Predictor variable names used in each CDI were also identified from each CDI’s MDCalc page. The predictor variables were grouped by mode of collection. Age, sex, weight, BMI, and race and ethnicity were considered *patient-level* variables. Nausea/vomiting, abdominal pain, fever, etc. were considered *signs and symptoms* variables. Vital signs and Glasgow Coma Scale score were considered *physical examination findings*.

Due to variability in predictor variable nomenclature between CDIs measuring identical values (e.g. “Systolic Blood Pressure” and “sBP”), naming modifications were applied upon data extraction to rename these predictor variables to match each other (e.g. “Systolic Blood Pressure” → Systolic BP; “sBP” → Systolic BP). All nomenclature modifications are displayed in Table A3. We defined other variables that may cause bias, such as those related to pain (e.g. Abdominal Pain) and family history. Family history is defined by the presence of chronic illness within the participant’s family resulting in a factor within the CDI’s algorithm calculation. CDIs with biased predictor variables, such as family history and race and ethnicity, were grouped and shown in Tables A1 and A2.

### Outcome variables

Outcome definitions and measures were manually identified from the text of each study. Outcome definitions describe the intended purpose or measurement of the CDI. These goals aim to include recommendations of medical treatment, diagnostic measure, or prognosis upon participant diagnosis. Outcome measures are defined as the benchmark measures used during the CDI’s development to test CDI internal validity. Recorded outcome measures include gold standard measures, previously validated measures from past literature, and/or reference measures. Outcome measures were not recorded if primary literature was a review, meta-analyses without explicit outcome measure description, or a descriptive article.

Determining inclusion of follow-up methodology as part of the outcome definition was conducted by assessing CDI study methodology for the implementation of recording data between at least 2 time points. Keywords assessed in methodology include *follow up, follow, database, dataset, phone*, etc. Direct follow-up was defined by in-person or telephone follow up specifically for participation within study outcome. Indirect follow-up was defined by researcher usage of databases or registries to identify patient data or health outcomes. Prospective observational studies were determined to be indirect follow-up. Primary literature that were meta-analyses were excluded from this analysis due to differences in study methods within these articles. At least 2 study team members separately reviewed each outcome measure to determine if follow-up was involved. A third study team member reconciled any conflicting determination of follow-up.

## Results

### Overview and characteristics of analyzed CDIs

We conducted a comprehensive analysis of CDIs by collecting key data for all CDIs available on the MDCalc website as of March 17, 2023. This process yielded a dataset of 690 CDIs, categorized by disease, system, purpose, chief complaint, and specialty according to MDCalc’s classification (Table 1). The CDIs included in the analysis exhibited diversity across several relevant characteristics. Most CDIs are fairly recent (median publication year 2017; see Fig. 1a), although the earliest publication in our data dates back to 1917. The number of new CDIs added each year has generally increased over time, with a recent decline corresponding to CDIs awaiting validation for inclusion in MDCalc. The number of predictor variables in the CDIs varied, with a median of 5 and a maximum of 48 (Fig. 1b). Additionally, the size of the development cohorts used to develop the CDIs varied widely (minimum 5, median 853, maximum 490,768; see Fig. 1c).

**Table 1.**
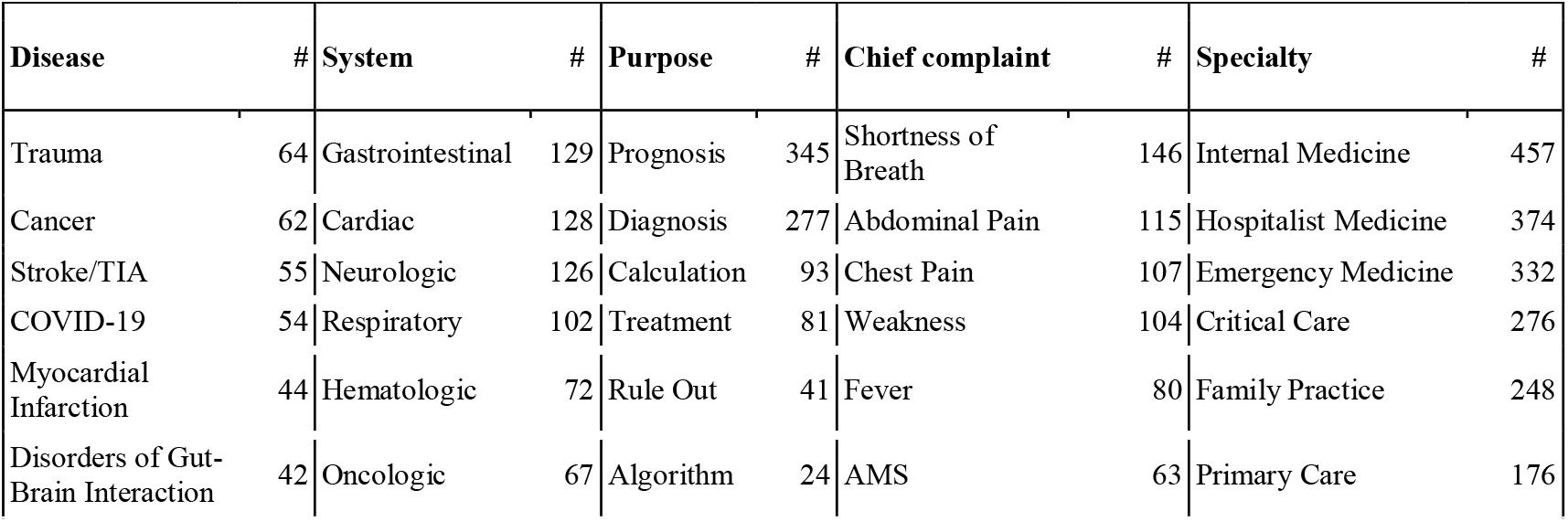
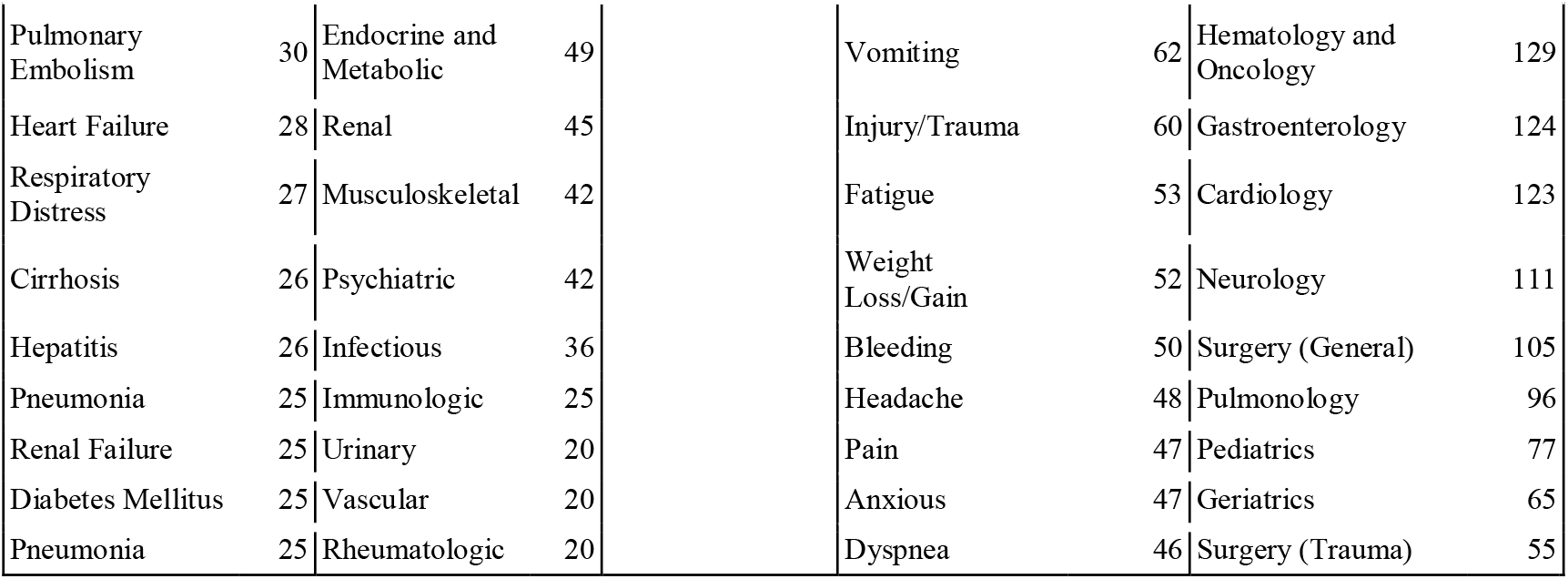
Categorizing CDIs by MDCalc’s classification system. 690 CDIs classified by disease, system, purpose, chief complaint, and specialty; only the top 10 most frequent values for each column are shown. CDIs may be labeled with multiple classes for a given categorization, e.g. a CDI can be categorized both under *Disease: Respiratory Distress* and *Disease: Pneumonia*). Abbreviations: *TIA: Transient ischemic attack, AMS: Altered mental status*.

**Fig. 1.**
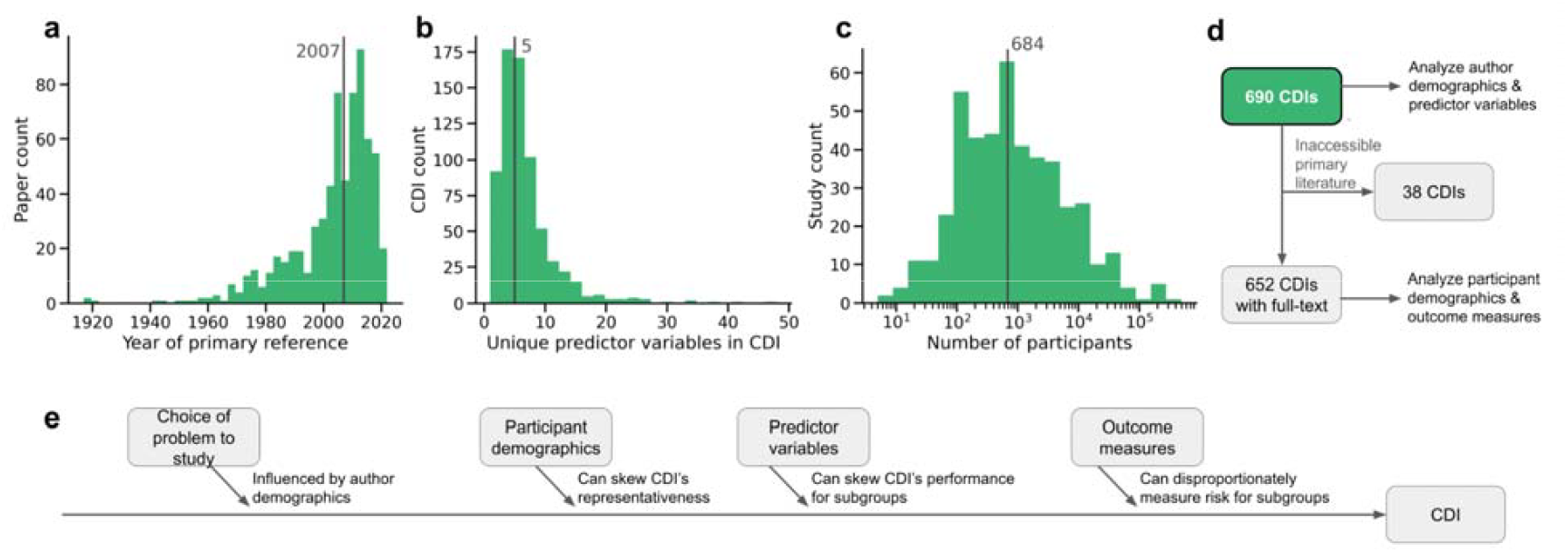
Overview of analyzed CDIs and key characteristics. The CDIs demonstrate diversity across several characteristics, including publication year (a), number of predictor variables (b), and size of the development cohort (c). Vertical lines represent the median values for each histogram. (d) A total of 690 CDIs were analyzed; 652 of these studies yield a full-text primary reference. (e) For each CDI, we analyze four factors that may introduce bias during CDI development.

To further analyze these 690 CDIs, we collected the full-text of the primary study whenever it is available, resulting in 652 CDIs with primary text (Fig. 1d); the 38 CDIs without accessible primary development literature are shown in Table A7. In what follows, we analyze four factors that may introduce bias during CDI development (Fig. 1e).

### Participant demographics

We evaluated the demographics of participant cohorts used in CDI development to assess potential implicit bias. Fig. 2a compares the racial and ethnic distributions of CDI participant cohorts with the U.S. population in 2020^20^. Note that different CDIs may be suitable for different populations, but as these are not marked consistently in the manuscripts or on MDCalc, we compare participant data to US population demographics. CDI cohorts significantly over-represent White-only participants (*p*=0.012) and significantly under-represent Latino populations (*p*=0.002). All *p*-values were calculated using 1-sample t-test with Benjamini-Hochberg false discovery rate correction^21^.

**Fig. 2.**
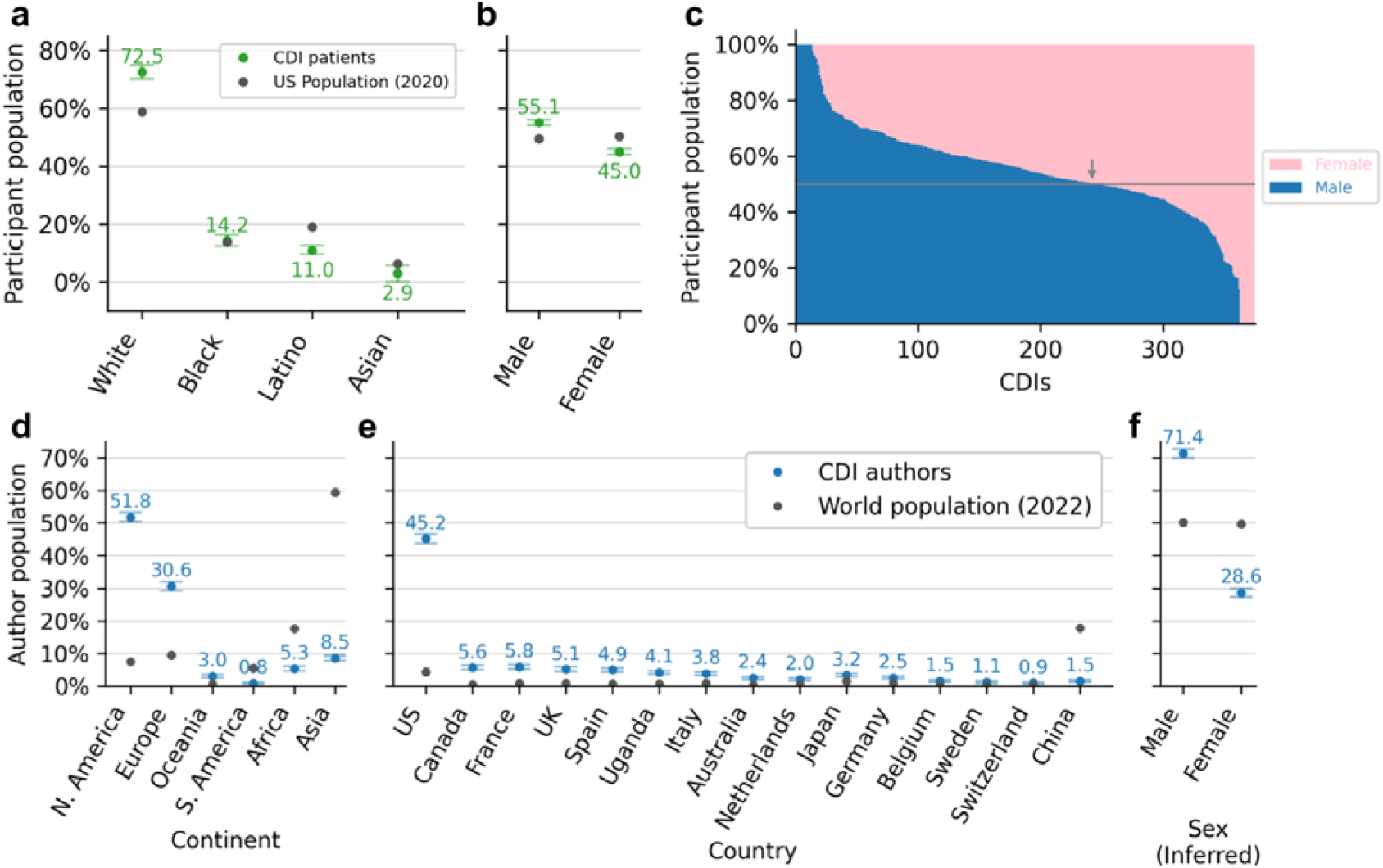
Demographics (self-reported) of CDI development participants and study authors. (a) Race and Ethnicity distribution of CDI development cohort participants compared to the US population across four racial categories; White, Black (or African American), Latino (or Hispanic), and Asian (or Pacific Islander). (b) Sex. (c) Sex breakdown (d) continent, (e) country, and (f) inferred sex. All values are sorted by the difference between the CDI author population and world population. Error values plot the medium over CDI primary studies (to avoid outlier effects) and error bars show standard error of the mean. Each country not shown accounts for less than 1% of CDI authors.

We further examined the sex distribution of participants in CDI development cohorts (Fig. 2b) and found a significant over–representation of male participants compared to female participants (*p*<10^−5^). As shown in Fig. 2c, 65% (242/374) of CDIs had a higher proportion of male participants than female participants. Additionally, 25 CDIs included participants of only one sex: 13 comprised exclusively male participants, and 12 comprised exclusively female participants. While most of these single-sex cohorts were for sex-specific conditions (e.g. testicular torsion^22^ or ovarian cancer^23^), some were not (e.g. HAM-D scale for depression^24^); see Table A4 for a list of all CDIs with single-sex cohorts.

### Author demographics

In a secondary analysis, we quantified potential biases in the author population of primary CDI studies. Specifically, we identified 3,819 author names and 1,234 author affiliations across 690 CDI studies and compared their demographics to the worldwide demographic distributions.^25^ Fig. 2d,e shows the distribution of author affiliations by continent or country. Authors are overwhelmingly in North America (52%) or Europe (31%) and nearly half of all authors are in the US (45%); all these discrepancies are significantly greater than the world population baseline (*p*<10^−5^). Fig. 2f further shows that sex distributions are skewed male even more than the participant population (71%), drastically more than worldwide demographics^26^ (*p*<10^−5^). These disparities are consistent with author demographics trends seen across medical publications beyond CDIs.^27,28^ While author demographics may not directly introduce bias, they may influence factors such as which outcomes are studied and which patient population is enrolled.

### Predictor Variables

We further investigated predictor variables that may be susceptible to introducing bias within CDIs, i.e. unintentionally providing less accurate or effective guidance for patients in particular subgroups. However, note that any of these predictor variables may not introduce bias, or may even correct for unintentional biases if used carefully.^29^

Fig 3a shows the most frequently used predictor variables in the MDCalc CDIs. We identify three common predictor variables which may be susceptible to introducing bias. First, *Abdominal pain* (present in 1.9% of CDIs) may be susceptible to clinical misinterpretation when severity is coded by a clinician rather than patient. Second, *Race and Ethnicity* (present in 1.9% of CDIs) may inadvertently skew predictions for racial groups, depending on its usage during CDI development. Third, *Family History* (present in 1.4% of CDIs) may indirectly be influenced by past familial medical knowledge, leading to differing impacts. We list the CDIs that explicitly use these predictor variables in Tables A1 and A2. The most common variable is Age (present in 32.6% of CDIs); we find that many of the most frequent predictor variables, e.g. *Sex*, tend to not contribute significant influence to the CDI to determine its outcome measure (see Fig A1). Quantifying this frequency helps identify variables that may be more important to study when understanding potential bias introduced by the variables within the CDI algorithm.

**Fig. 3.**
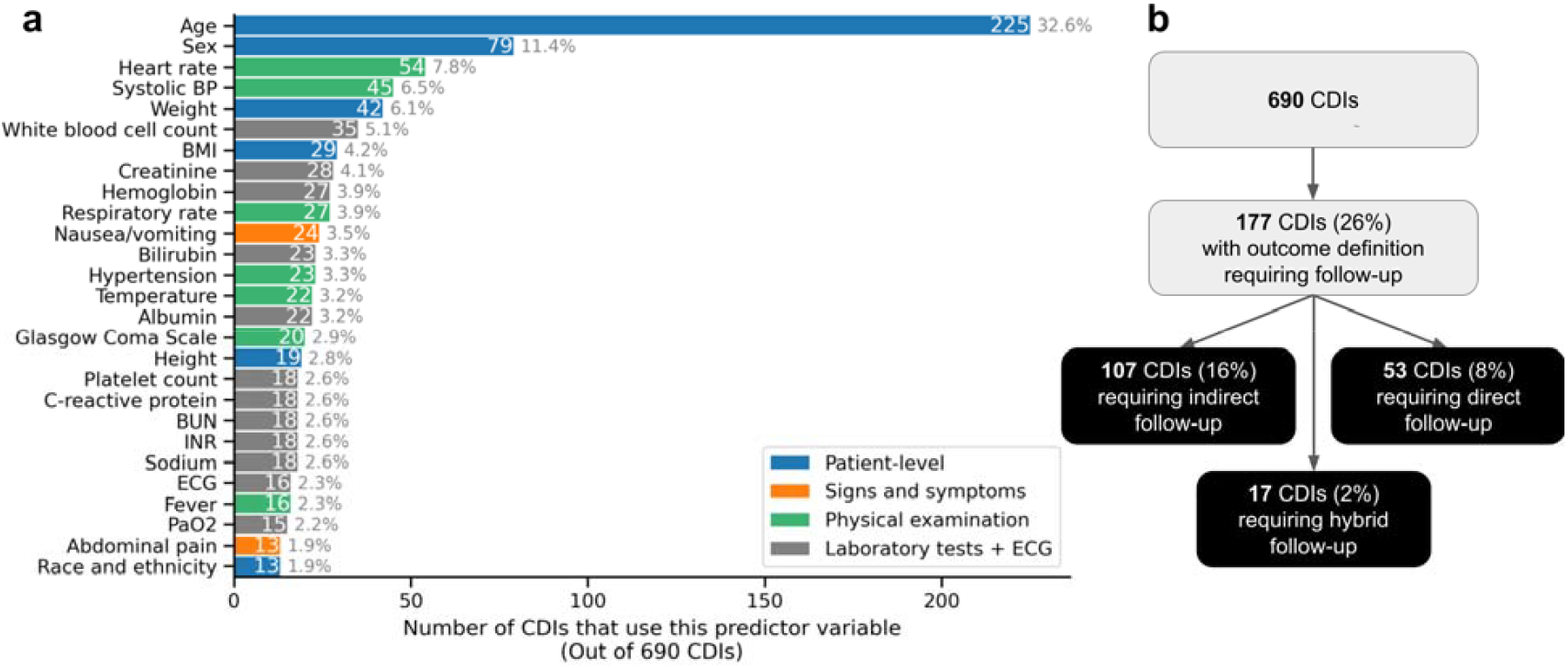
Frequency of predictor variables in MDCalc CDIs. Colors represent different categories of predictor variables dependent on type of collection. Blue represents participant-level predictor variables. Orange represents signs and symptoms upon participant presentation. Green represents physical examination findings. Gray represents laboratory tests including ECG. *Abbreviations*. BP: *Blood pressure*, BMI: *Body mass index*, BUN: *Blood urea nitrogen*, INR: *International normalized ratio blood test*, ECG: *electrocardiogram*. Variables appearing in less than 13 CDIs are not shown.

### Outcome measures

Finally, we investigated the outcome definition itself as a source of CDI bias. While there are many potential factors of the outcome definition, we specifically focused on the presence of *follow-up* in the outcome definition. Access to follow-up can vary by subgroup due to socioeconomic constraints^30^, which can lead to differential verification bias. For example, the Canadian CT Head Rule^31^, excluded patient data from analysis if investigators could not reach patients for follow-up. We define follow-up as a continuous recording of health data following an initial care visit. Follow-up can be “direct”, described as telephone or in-person visits, or “indirect”, described by health information present in a database. This factor potentially introduces bias, as wealthier participants are more likely to receive follow-up and meet the criteria for the outcome definition. We manually identified 177 CDIs (26%) that involve follow-up as part of the primary outcome definition (Fig 3b). Of these, 53 require direct follow-up and 17 require hybrid follow-up (together 10% of CDIs), both categories that are more prone to bias than indirect follow-up. See Table A6 for a full list of these CDIs involving follow-up.

## Discussion

Our study analyzes 690 MDCalc CDIs and provides the first quantitative meta-analysis of bias in CDIs. The work here adds to a growing literature that finds broad evidence for implicit bias in data-driven clinical algorithms and recommends improvements in best practices for CDI development.^2,32–36^ We now briefly discuss the four factors related to CDI development we analyzed along with concrete takeaways for developers and users of CDIs.

### Participant and investigator demographics

The over-representation of male participants in developing CDIs has various explanations and may raise concerns about the broader applicability of particular CDIs. The male skew may be rooted in past Western regulatory policies advising the exclusion of women of childbearing potential from studies.^37^ This skew may have persisted past these policies, as investigators sought to maintain consistency with prior male-only clinical trials.^38,39^ Today, quantifying this skew can be challenging, as literature often fails to report results by sex and other demographics, even after guidance and mandates from the NIH.^40,41^ Going beyond enrollment, many studies may specifically focus on male-specific health issues, e.g. testicular torsion and prostate cancer. Therefore, we recommend that bias mitigation starts as early as designing the enrollment process for participants to optimize the generalizability of the CDI.

The demographic and geographic skews we observed in CDI development (Fig 2) can harm the global generalizability of CDIs. Specifically, CDIs are largely developed in Western countries and often used in non-Western countries with differing populations and disease profiles.^42^ For example, the Kruis Score for diagnosing irritable bowel syndrome (IBS) enrolled a development cohort that overrepresented women, which is consistent with the higher prevalence of IBS in women in Western countries.^43^ However, IBS prevalence has been shown to be equally common between males and females in Asia, highlighting how risk factors can be limited by the regional demographics of a study population.^44^ To mitigate the effects of these demographic skews, we recommend that CDI developers should disclose awareness of risk factors for disease in their methodology for enrolling participants to highlight the limited generalizability of their instrument. Moreover, to reduce potential harm in CDI usage, validation studies should be conducted to test CDIs on underrepresented populations of the development cohort to assess generalizability prior to being officially approved for use in clinical settings.

### Predictor variables

The inclusion of predictor variables which are susceptible to bias is nuanced, and therefore should be treated with caution. For example, *Race* as a predictor variable has been subject to a great deal of scrutiny,^45,7,8^ even leading to modern revisions for some instruments without Race, e.g. in eGFR measurement for kidney function^46^ or prediction of vaginal birth after cesarean delivery^47^. However, these same race adjustments, if used correctly in a CDI, may help correct for racial disparities in data quality.^48^ CDI developers and clinicians should be aware of the potential bias introduced through race variables (in line with MDCalc’s Statement on Race^49^). Going beyond Race, CDIs should communicate to clinicians the biases that particular predictor variables may be susceptible to introducing, e.g. clinician perceptions of pain.

### Outcome measures

Outcome measures are a prevalent and often understudied source of bias in clinical algorithms^30^. We limited our analysis to studying *follow-up* (both direct and indirect) as an alternative reference standard for the primary outcome. However, there may be systemic implicit bias related to follow-up procedures that unintentionally limit the types of participants to those who can comply with follow–up methodology for the primary outcome. Direct follow-up poses concern for bias as in-person visits and telephone communication can be limited by transportation and phone accessibility, low-income status, physical disability, and language barriers. To deal with this, some analyzed CDI development studies needed to either exclude patient data (e.g. HEART Score^50^) or input assumed data (e.g. Modified NIH Stroke Scale/Score^51^) to retain appropriate presence within the study dataset for patients who lack follow-up data. Indirect follow-up raises different concerns with the wide usage of research databases within CDI development. While these databases often hold large sets of patient information, they are subject to biases that include missing data, sample size and underestimation, and measurement errors.^32^ Switching clinicians and areas of living may lead to their data being stored in multiple electronic databases that do not communicate with each other. CDI developers must remain aware of data fragmentation among patients with transient living situations.

### Limitations

Our study has several important limitations. First, we only analyzed primary literature within MDCalc, which may not provide a perfectly accurate representation of the international landscape of CDI development and usage. Moreover, we perform analysis using metadata but do not look directly at underlying data and algorithms used to develop CDIs. It is possible that direct or indirect follow-up was included in CDI studies but were not discussed or specified within the study methods. We compared CDI participant data to US population data because it was difficult to get worldwide data on racial and ethnic demographics. However, it is clear that disparities from the representation of minority groups are exacerbated when comparing CDI populations to worldwide populations (e.g. White over-representation and Asian under-representation). Bias can manifest itself in many ways beyond the components of CDI development that we study here including the validation and formulation phases. Future research should explore other potential signals of bias such as subjective clinician assessments and the use of costly diagnostic tests in CDI development.

### Concrete recommendations

In conclusion, we found CDIs may be biased at the patient, author, predictor, and outcome level. We recommend that implicit bias is taken into consideration by all parties involved in CDI usage: (i) investigators developing and validating CDIs, (ii) services implementing CDIs, e.g. MDCalc, and (iii) clinicians using CDIs. Investigators creating CDIs must consider and report numerous factors, including the demographics of their enrollment cohort and their predictor/outcome variables. Once a CDI has been developed, the addition of model cards can be an effective strategy to showcase the CDI model for describing its benefits (intended uses, target population, and efficacy evaluations) and limitations (risks, inappropriate settings, and generalizability).^15^ While CDIs hold great potential for enhancing patient care through data-driven standardization, our findings underscore the need for careful consideration of bias in their development. By addressing these biases, we can work towards the development of more equitable and effective CDIs, ultimately contributing to more inclusive healthcare systems. Future research should continue to explore and quantify potential roots of bias in CDI development, and work towards developing strategies to mitigate these biases.

## Data Availability

All data and code for reproducing the results in this manuscript is made publicly available through MDCalc and processed data is made available at https://github.com/csinva/clinical-rule-analysis.

## Code Availability

All code for reproducing the results in this manuscript is made publicly available at https://github.com/csinva/clinical-rule-analysis.

## Author Contributions

Concept and design: JKO, CS, ZO, AK; Acquisition, analysis or interpretation of data: JKO, CS, KW, AK; Draft manuscript: JKO, CS, KW, JF, ZO, AK; Statistical Analysis: CS; Supervision: CS, AK. All authors contributed to the analysis of results, reviewed this manuscript, and revised it critically for important intellectual content and approved of the version as submitted.

## Acknowledgements

The authors declare no conflicts of interest exist. We thank the MDCalc team for creating a valuable resource for organizing CDIs.

## References

1. Reilly, B. M. & Evans, A. T. Translating clinical research into clinical practice: impact of using prediction rules to make decisions. Ann. Intern. Med. 144, 201–209 (2006).

2. Stiell, I. G. & Bennett, C. Implementation of Clinical Decision Rules in the Emergency Department. Acad. Emerg. Med. 14, 955–959 (2007).

3. Paulus, J. K. & Kent, D. M. Predictably unequal: understanding and addressing concerns that algorithmic clinical prediction may increase health disparities. Npj Digit. Med. 3, 1–8 (2020).

4. Obermeyer, Z., Powers, B., Vogeli, C. & Mullainathan, S. Dissecting racial bias in an algorithm used to manage the health of populations. Science 366, 447–453 (2019).

5. Mauro, M. et al. A scoping review of guidelines for the use of race, ethnicity, and ancestry reveals widespread consensus but also points of ongoing disagreement. Am. J. Hum. Genet. 109, 2110 (2022).

6. Callier, S. L. The Use of Racial Categories in Precision Medicine Research. Ethn. Dis. 29, 651 (2019).

7. Vyas, D. A., Eisenstein, L. G. & Jones, D. S. Hidden in plain sight—reconsidering the use of race correction in clinical algorithms. New England Journal of Medicine vol. 383 874–882 (2020).

8. Perez-Rodriguez, J. & de la Fuente, A. Now is the Time for a Postracial Medicine: Biomedical Research, the National Institutes of Health, and the Perpetuation of Scientific Racism. Am. J. Bioeth. AJOB 17, 36–47 (2017).

9. Ozanne, E. M. et al. Bias in the Reporting of Family History: Implications for Clinical Care. J. Genet. Couns. 21, 547–556 (2012).

10. Howe, C. J., Cole, S. R., Lau, B., Napravnik, S. & Eron, J. J. Selection Bias Due to Loss to Follow Up in Cohort Studies. Epidemiol. Camb. Mass 27, 91–97 (2016).

11. Syed, S. T., Gerber, B. S. & Sharp, L. K. Traveling towards disease: transportation barriers to health care access. J. Community Health 38, 976–993 (2013).

12. Love, H. B., Stephens, A., Fosdick, B. K., Tofany, E. & Fisher, E. R. The impact of gender diversity on scientific research teams: a need to broaden and accelerate future research. Humanit. Soc. Sci. Commun. 9, 1–12 (2022).

13. MDCalc - Medical calculators, equations, scores, and guidelines. MDCalc https://www.mdcalc.com/.

14. Collins, G. S., Reitsma, J. B., Altman, D. G. & Moons, K. G. Transparent reporting of a multivariable prediction model for individual prognosis or diagnosis (TRIPOD): the TRIPOD Statement. BMC Med. 13, 1 (2015).

15. Sendak, M. P., Gao, M., Brajer, N. & Balu, S. Presenting machine learning model information to clinical end users with model facts labels. NPJ Digit. Med. 3, 41 (2020).

16. Page, M. J. et al. The PRISMA 2020 statement: an updated guideline for reporting systematic reviews. Syst. Rev. 10, 89 (2021).

17. Clayton, J. A. & Tannenbaum, C. Reporting Sex, Gender, or Both in Clinical Research? JAMA 316, 1863–1864 (2016).

18. Flanagin, A., Frey, T., Christiansen, S. L., & AMA Manual of Style Committee. Updated Guidance on the Reporting of Race and Ethnicity in Medical and Science Journals. JAMA 326, 621–627 (2021).

19. Ouyang, L. et al. Training language models to follow instructions with human feedback. Preprint at 10.48550/arXiv.2203.02155 (2022).

20. Bureau, U. C. Census.gov | U.S. Census Bureau Homepage. Census.gov https://www.census.gov/en.html.

21. Benjamini, Y. & Hochberg, Y. Controlling the False Discovery Rate: A Practical and Powerful Approach to Multiple Testing. J. R. Stat. Soc. Ser. B Methodol. 57, 289–300 (1995).

22. Barbosa, J. A. et al. Development and initial validation of a scoring system to diagnose testicular torsion in children. J. Urol. 189, 1859–1864 (2013).

23. Jacobs, I. et al. A risk of malignancy index incorporating CA 125, ultrasound and menopausal status for the accurate preoperative diagnosis of ovarian cancer. Br. J. Obstet. Gynaecol. 97, 922–929 (1990).

24. Hamilton, M. A rating scale for depression. J. Neurol. Neurosurg. Psychiatry 23, 56–62 (1960).

25. World Population Prospects 2022: Summary of Results | Population Division. https://www.un.org/development/desa/pd/content/World-Population-Prospects-2022.

26. The CIA World Factbook 2021-2022. Skyhorse Publishing https://www.skyhorsepublishing.com/9781510763814/the-cia-world-factbook-2021-2022.

27. Charpignon, M.-L. et al. Does diversity beget diversity? A scientometric analysis of over 150,000 studies and 49,000 authors published in high-impact medical journals between 2007 and 2022. MedRxiv Prepr. Serv. Health Sci. 2024.03.21.24304695 (2024) doi:10.1101/2024.03.21.24304695.

28. Saloojee, H. & Pettifor, J. M. Maximizing Access and Minimizing Barriers to Research in Low- and Middle-Income Countries: Open Access and Health Equity. Calcif. Tissue Int. 114, 83 (2023).

29. Zink, A., Obermeyer, Z. & Pierson, E. Race Corrections in Clinical Models: Examining Family History and Cancer Risk. medRxiv 2023–03 (2023).

30. Yan, M., Pencina, M. J., Boulware, L. E. & Goldstein, B. A. Observability and its impact on differential bias for clinical prediction models. J. Am. Med. Inform. Assoc. 29, 937–943 (2022).

31. Stiell, I. G. et al. The Canadian CT Head Rule for patients with minor head injury. The Lancet 357, 1391–1396 (2001).

32. Gianfrancesco, M. A., Tamang, S., Yazdany, J. & Schmajuk, G. Potential Biases in Machine Learning Algorithms Using Electronic Health Record Data. JAMA Intern. Med. 178, 1544–1547 (2018).

33. Heerink, J. S., Oudega, R., Hopstaken, R., Koffijberg, H. & Kusters, R. Clinical decision rules in primary care: necessary investments for sustainable healthcare. Prim. Health Care Res. Dev. 24, e34 (2023).

34. Shapiro, S. E. Guidelines for developing and testing clinical decision rules. West. J. Nurs. Res. 28, 244–253 (2006).

35. Stiell, I. G. & Wells, G. A. Methodologic standards for the development of clinical decision rules in emergency medicine. Ann. Emerg. Med. 33, 437–447 (1999).

36. Ebell, M. AHRQ White Paper: Use of clinical decision rules for point-of-care decision support. Med. Decis. Mak. Int. J. Soc. Med. Decis. Mak. 30, 712–721 (2010).

37. Merone, L., Tsey, K., Russell, D. & Nagle, C. Sex Inequalities in Medical Research: A Systematic Scoping Review of the Literature. Womens Health Rep. 3, 49 (2022).

38. Holdcroft, A. Gender bias in research: how does it affect evidence based medicine? J. R. Soc. Med. 100, 2 (2007).

39. Lee, E. & Wen, P. Gender and sex disparity in cancer trials. ESMO Open 5, e000773 (2020).

40. Poon, R. et al. Participation of Women and Sex Analyses in Late-Phase Clinical Trials of New Molecular Entity Drugs and Biologics Approved by the FDA in 2007–2009. J. Womens Health 22, 604 (2013).

41. Geller, S. E. et al. The More Things Change, the More They Stay the Same: A Study to Evaluate Compliance With Inclusion and Assessment of Women and Minorities in Randomized Controlled Trials. Acad. Med. J. Assoc. Am. Med. Coll. 93, 630–635 (2018).

42. Peiffer-Smadja, N. et al. Paving the Way for the Implementation of a Decision Support System for Antibiotic Prescribing in Primary Care in West Africa: Preimplementation and Co-Design Workshop With Physicians. J. Med. Internet Res. 22, e17940 (2020).

43. Lovell, R. M. & Ford, A. C. Effect of gender on prevalence of irritable bowel syndrome in the community: systematic review and meta-analysis. Am. J. Gastroenterol. 107, 991–1000 (2012).

44. Gwee, K. A. et al. Second Asian Consensus on Irritable Bowel Syndrome. J. Neurogastroenterol. Vienna AustriaMotil. 25, 343 (2019).

45. Suriyakumar, V., Ghassemi, M. & Ustun, B. When Personalization Harms: Reconsidering the Use of Group Attributes in Prediction. (2022). doi:10.48550/arXiv.2206.02058.

46. Eneanya, N. D., Yang, W. & Reese, P. P. Reconsidering the Consequences of Using Race to Estimate Kidney Function. JAMA 322, 113–114 (2019).

47. Grobman, W. A. et al. Prediction of vaginal birth after cesarean delivery in term gestations: a calculator without race and ethnicity. Am. J. Obstet. Gynecol. 225, 664.e1–664.e7 (2021).

48. Zink, A., Obermeyer, Z. & Pierson, E. Race Corrections in Clinical Models: Examining Family History and Cancer Risk. 2023.03.31.23287926 Preprint at 10.1101/2023.03.31.23287926 (2023).

49. MDCalc Statement on Race. MDCalc https://www.mdcalc.com/race.

50. Six, A. J., Backus, B. E. & Kelder, J. C. Chest pain in the emergency room: value of the HEART score. Neth. Heart J. Mon. J. Neth. Soc. Cardiol. Neth. Heart Found. 16, 191–196 (2008).

51. Lyden, P. D. et al. A modified National Institutes of Health Stroke Scale for use in stroke clinical trials: preliminary reliability and validity. Stroke 32, 1310–1317 (2001).

