## Supplemental Information for "Systematic Bias in Clinical Decision Instrument Development: A Quantitative Meta-Analysis"

### Abstract

Clinical decision instruments (CDIs) face an equity dilemma. On the one hand, they often reduce disparities in patient care through data-driven standardization of best practices. On the other hand, this standardization may itself inadvertently perpetuate bias and inequality within healthcare systems. Here, we quantify different measures of potential for implicit bias present in CDI development that can inform future CDI development. We find evidence for systematic bias in the development of 690 CDIs that underwent validation through various analyses: self-reported participant demographics are skewed—e.g. 73% of participants are White, 55% are male; investigator teams are geographically skewed—e.g. 52% in North America, 31% in Europe; CDIs use predictor variables that may be prone to bias—e.g. 13 CDIs explicitly use *Race and Ethnicity*; outcome definitions may further introduce bias—e.g. 28% of CDIs involve follow-up, which may disproportionately skew outcome representation based on socioeconomic status. As CDIs become increasingly prominent in medicine, we recommend that these factors are considered during development and clearly conveyed to clinicians using CDIs.

### Supplementary Information

Some predictor variables may be very frequent, but nevertheless relatively unimportant in CDIs. Fig 6 investigates the relative importance of frequent variables in CDIs. It analyzes scoring CDIs which assign points based on predictor variables; this excludes other types of CDIs, e.g. CDIs that use nonlinear formulas. We calculate variable importance as the fraction of total points. For example, if a CDI assigns 1 point for variable A and 3 points for variable B, then variable A receives a variable importance of ¼ and variable B receives a variable importance of ¾. Some very frequent variables (e.g. Sex, BMI) tend to have very low importance whereas some less frequent variables (e.g. C-reactive protein, BUN), tend to have higher importance.

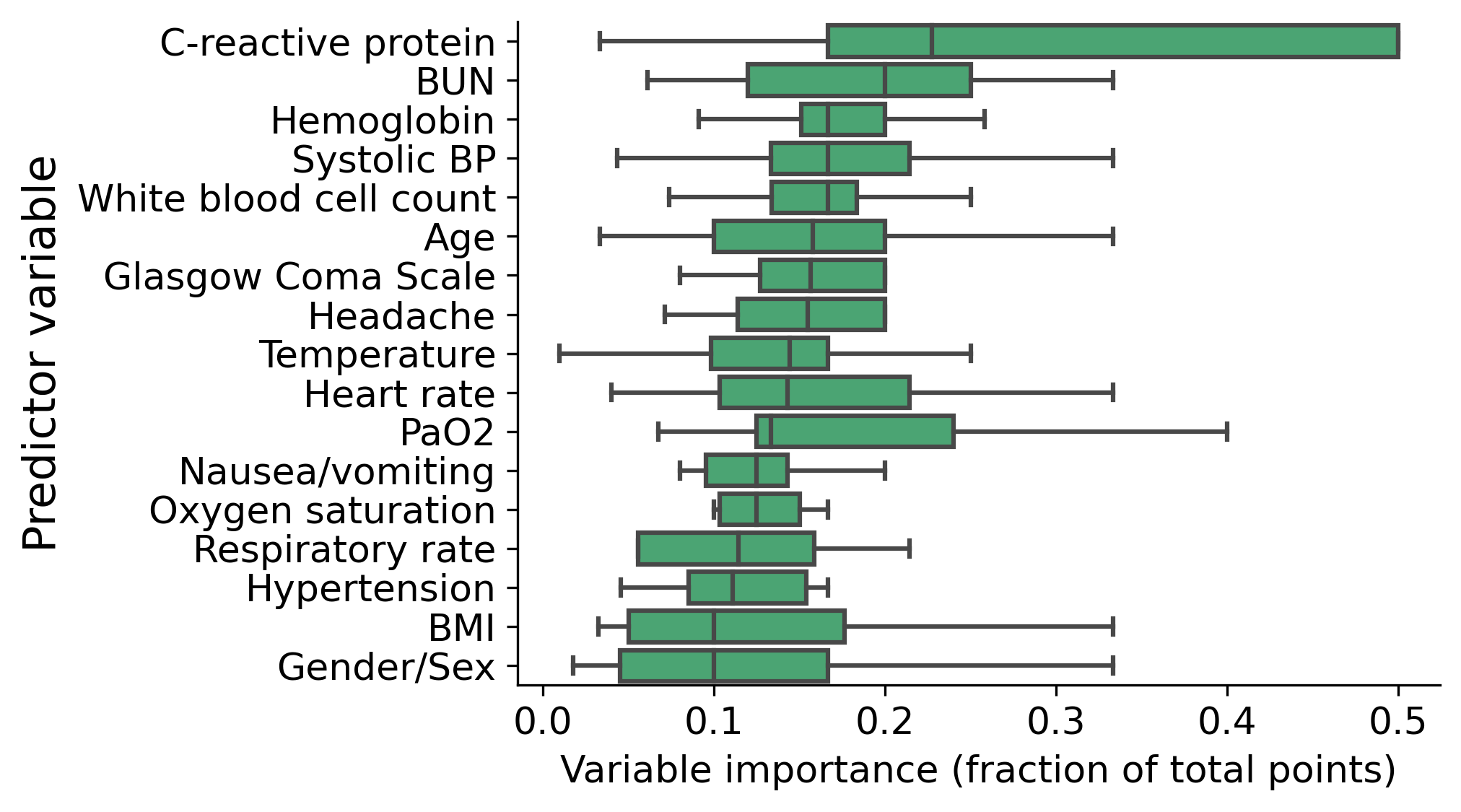

**Fig A1 | Variable importance of frequent predictor variables in CDIs.** Restricted to scoring CDIS (309 / 690 CDIs) and to variables which appear in at least 9 scoring CDIs. Each box shows the median, interquartile range, and extended lines show 1.5 of the interquartile range. *Abbreviations. BP: Blood pressure, BMI: Body mass index, BUN: Blood urea nitrogen.*

**Table A1 |** CDIs including *Race and Ethnicity* predictor variables.

| **CDI** | **Usage description** | **Race variable** |
| --- | --- | --- |
| ASCVD (Atherosclerotic Cardiovascular Disease) 2013 Risk Calculator from AHA/ACC | 10-year heart disease or stroke risk. | Race |
| ASCVD (Atherosclerotic Cardiovascular Disease) Risk Algorithm including Known ASCVD from AHA/ACC | 10-year heart disease or stroke risk and statin recommendations. | Race |
| Australian Type 2 Diabetes Risk (AUSDRISK) Assessment Tool | Estimates diabetes risk. | Ethnicity |
| CKD-EPI Equations for Glomerular Filtration Rate (GFR) | Estimates GFR. | Race |
| COVID-19 Inpatient Risk Calculator (CIRC) | Inpatient COVID mortality or severe disease progression. | White |
| Denver HIV Risk Score | HIV risk screening. | Race and ethnicity |
| Estimated/Expected Peak Expiratory Flow (Peak Flow) | Quantifies asthma severity. | Race and ethnicity |
| GWTG-Heart Failure Risk Score | Predicts in-hospital heart failure mortality. | Black race |
| Gail Model for Breast Cancer Risk | Breast CA risk based on demographic and clinical data. | Asian-American sub race, Race and ethnicity |
| Kinetic Estimated Glomerular Filtration Rate (keGFR) | keGFR estimation. | Black race |
| Licurse Score for Renal Ultrasound | Hydronephrosis probability. | Race |
| MDRD GFR Equation | For CKD patients (not for AKI). | Black race |
| STONE Score for Uncomplicated Ureteral Stone | Predicts ureteral stones, before ultrasound. | Race |

**Table A2 |** CDIs including “Family history” variables.

| **CDI** | **Usage description** | **Family history variables** |
| --- | --- | --- |
| ACC/AHA Heart Failure Staging | Heart failure staging and therapy recommendations. | Patient with family history of cardiomyopathy |
| ASAS Criteria for Axial Spondyloarthritis (SpA) | Axial SpA diagnosis. | Family history of SpA |
| ASAS Criteria for Peripheral SpondyloArthritis (SpA) | Peripheral SpA diagnosis. | Positive family history of SpA |
| CASPAR Criteria for Psoriatic Arthritis | Psoriatic arthritis diagnosis. | Current psoriasis, personal history of psoriasis, or family history of psoriasis |
| Cambridge Diabetes Risk Score | T2DM risk. | Family history |
| Caprini Score for Venous Thromboembolism (2005) | Stratifies risk of VTE in surgical patients. | Family history of thrombosis |
| Dutch Criteria for Familial Hypercholesterolemia (FH) | Diagnoses familial hypercholesterolemia. | Patient has elevated cholesterol, family history of FH, and/or family history of premature cardiac death |
| FINDRISC (Finnish Diabetes Risk Score) | T2DM risk. | Family history of diabetes |
| Opioid Risk Tool (ORT) for Narcotic Abuse | Risk of narcotic abuse/misuse. | Family history of illegal drug abuse, Family history of prescription drug abuse, Family history of alcohol abuse |
| Simon Broome Diagnostic Criteria for Familial Hypercholesterolemia (FH) | Diagnoses familial hypercholesterolemia (FH) | Family history of premature CVD events, Family history of extremely high cholesterol |

**Table A3 | Renaming rules for predictor variables.** Rules are applied starting from the left column: if a string contains a value in the *Case-insensitive match* column or the *Case-sensitive match* column, it is renamed to that value. Then, if a string starts with any of the prefixes in *Case-insensitive prefix match,* it is renamed to the prefix. Finally, any value matching a rule in *Case-sensitive rename* is renamed following the rule.

| **Case-insensitive match** | **Case-sensitive match** | **Case-insensitive prefix match** | **Case-sensitive rename** |
| --- | --- | --- | --- |
| Ethnicity | ALT | Age | Systolic Blood Pressure → Systolic BP |
| D-dimer | AST | ASA | Ethnicity → Race and Ethnicity |
| Appetite | BMI | Albumin | White blood cell → White blood cell count |
| Level of consciousness | CRP | Anxiety | WBC → White blood cell count |
| Chest x-ray | C-reactive protein | Atrial fibrillation | CRP → C-reactive protein |
| Weight loss | CD4 | ED visits | Diminished breath sounds → Decreased breath sounds |
| Karnofsky | CHF | EKG | EKG → ECG |
| Dysarthria | ECG | ESR | Female → Sex |
| Facial palsy | EKG | Endoscopy | GCS → Glasgow Coma Scale |
| Acidosis | HDL | BMI | Glasgow Coma Score → Glasgow Coma Scale |
| Abdominal pain | GCS | BUN | Pulse → Heart rate |
| Altered mental status | WBC | Biliary | Immobilization → Immobilized |
| Bilirubin | INR | Calcium | Intubation → Intubated |
| Eosinophilia | LDL | Congestive heart failure | Leukocyte → White blood cell count |
| Enthesitis | LDH | Creatinine | Vomiting → Nausea/vomiting |
| White blood cell | NIHSS | Dementia | NIHSS → NIH Stroke Scale |
| Erythrocyte sedimentation rate | NYHA | Distracting | Obesity → BMI |
| Estimated blood loss | PSA | ECOG | O₂ sat → Oxygen saturation |
| Female | PaCO₂ | Erythema | PaO₂ → PaO2 |
| Ferritin | PaO₂ | Glucose | Patient age → Age |
| gestational age | PaO2 | Hematocrit | Patient sex → Sex |
| Glasgow Coma Scale | sBP | Hemoglobin | Persistent vomiting → Nausea/vomiting |
| Heart rate |  | Race | Platelet → Platelet count |
| Pulse |  | Regional lymph node | Pregnant → Pregnancy |
| Headache |  | Respiratory rate | SpO₂ → Oxygen saturation |
| Height |  | Scalp hematoma | Sp02 → Oxygen saturation |
| Hematuria |  | Sex | Suicid → Suicidality |
| Fever |  | Systolic BP | Temp → Temperature |
| Blood in stool |  | Male | Tremor → Tremors |
| Diastolic BP |  | Nausea | Triglyceride → Triglycerides |
| Hypotension |  | Oxygen saturation | sBP → Systolic BP |
| Hypertension |  | Platelet |  |
| Immobilized |  | Potassium | Diastolic blood pressure → Diastolic BP |
| Immobilization |  | Pregnancy | Systolic pressure → Systolic BP |
| Insulin |  | Pregnant | Nausea → Nausea/vomiting |
| Intraventricular hemorrhage |  | SaO₂ | White → Race and Ethnicity |
| Intubation |  | Seizure | Race → Race and Ethnicity |
| Intubated |  | Sodium | Gender → Sex |
| Lactate |  | Sp02 | Male → Sex |
| Length of stay |  | SpO₂ |  |
| Leukocyte |  | Temp |  |
| Marked change in tone |  | Tremor |  |
| NIH Stroke Scale |  | Triglyceride |  |
| Age |  | Wheezing |  |
| (Age |  | eGFR |  |
| Verbal response |  | Weight |  |
| Diastolic blood pressure |  | Suidicid |  |
| Systolic pressure |  |  |  |
| Vomiting |  |  |  |

**Table A4 | CDIs which have development cohorts that entirely consist of patients of a single sex.** There are 13 male-only cohorts and 12 female-only cohorts. The mean fraction of male patients across all cohorts is 55.0%. When excluding single-sex cohorts, the mean fraction of male patients is 54.8%.

| **CDI** | **Description** | **# Male** | **# Female** |
| --- | --- | --- | --- |
| CKD Prediction in HIV+ Patients | 5 year likelihood of CKD in HIV. | 21590 | 0 |
| UCSF-CAPRA Score for Prostate Cancer Risk | Post-treatment outcomes for prostate cancer. | 10627 | 0 |
| Veterans Aging Cohort Study (VACS) 1.0 Index | All-cause mortality for patients with HIV or HCV. | 4932 | 0 |
| PSA Doubling Time (PSADT) Calculator | PSA survival. | 1997 | 0 |
| D'Amico Risk Classification for Prostate Cancer | Assesses 5 year failure of prostate CA treatment failure. | 1872 | 0 |
| Testicular Workup for Ischemia and Suspected Torsion (TWIST) | Testicular torsion diagnosis. | 338 | 0 |
| Bastion Classification of Lower Limb Blast Injuries | Lower limb blast injury. | 103 | 0 |
| Prostate Tumor Volume & Density | Tumor volume and PSA density. | 100 | 0 |
| Hamilton Depression Rating Scale (HAM-D) | Depression severity. | 49 | 0 |
| Pain Assessment in Advanced Dementia Scale (PAINAD) | Pain level in dementia patients. | 19 | 0 |
| CEDOCS Score for Emergency Department Overcrowding | Estimates severity of overcrowding in community EDs. | 12 | 0 |
| Phenytoin (Dilantin) Correction for Albumin / Renal Failure | Corrected phenytoin level. | 9 | 0 |
| Stanford Sleepiness Scale | Quantifies sleepiness. | 5 | 0 |
| Gail Model for Breast Cancer Risk | Breast CA risk based on demographic and clinical data. | 0 | 284780 |
| Vaginal Birth After Cesarean (MFMU) | Successful vaginal birth prediction. | 0 | 11687 |
| VBAC Risk Score for Successful Vaginal Delivery (Flamm Model) | Quantifies likelihood of vaginal birth after previous C-section. | 0 | 5022 |
| Prevention and Incidence of Asthma and Mite Allergy (PIAMA) Risk Score | Risk of asthma for school age children. | 0 | 4146 |
| Modified Bishop Score for Vaginal Delivery and Induction of Labor | Likelihood of successful vaginal delivery. | 0 | 1189 |
| Charlson Comorbidity Index (CCI) | 10-year survival. | 0 | 685 |
| Danger Assessment Tool for Domestic Abuse | Death by intimate partner violence. | 0 | 666 |
| BWH Egg Freezing Counseling Tool (EFCT) | Predicts likelihood of live birth for elective egg freezing in women. | 0 | 520 |
| Humiliation, Afraid, Rape, Kick (HARK) | Primary care domestic abuse detection. | 0 | 232 |
| Hurt, Insult, Threaten, Scream (HITS) Score | Detects domestic abuse in healthcare settings. | 0 | 160 |
| Risk of Malignancy Index (RMI) for Ovarian Cancer | Ovarian cancer risk for adnexal mass. | 0 | 143 |
| Bristol Stool Form Scale | Stool classification. | 0 | 66 |

**Table A5 | Fraction of patient cohort identified as male, stratified by Disease, System, and Specialty.** Categories containing less than 5 CDIs are excluded from this table. Some specialties have understandable imbalances, e.g. CDIs related to OB-Gyn have predominantly female patients whereas CDIs related to Urology have predominantly male patients.

| **Disease** | | **System** | | | **Specialty** | |
| --- | --- | --- | --- | --- | --- | --- |
| Public Health | 0.28 | Reproductive |  | 0.29 | OB-Gyn | 0.19 |
| Rheumatoid Arthritis | 0.34 | Rheumatologic |  | 0.36 | Rheumatology | 0.36 |
| Depression/Suicidality | 0.42 | Any/All |  | 0.47 | Orthopedics | 0.42 |
| Pulmonary Embolism | 0.45 | Vascular |  | 0.48 | Oral and Maxillofacial Surgery | 0.47 |
| Diabetes Mellitus | 0.47 | Musculoskeletal |  | 0.48 | Allergy and Immunology | 0.47 |
| Deep Venous Thrombosis | 0.47 | Dermatologic |  | 0.51 | Radiation Oncology | 0.49 |
| Delirium | 0.50 | Psychiatric |  | 0.52 | Surgery (Vascular) | 0.49 |
| Sepsis | 0.51 | Respiratory |  | 0.54 | Psychiatry | 0.50 |
| Chronic Pain | 0.51 | Endocrine and Metabolic |  | 0.55 | Dermatology | 0.51 |
| Respiratory Failure | 0.51 | Cardiac |  | 0.56 | Family Practice | 0.51 |
| Anxiety | 0.51 | Hematologic |  | 0.56 | Endocrinology | 0.52 |
| Obesity | 0.51 | Neurologic |  | 0.57 | Neurosurgery | 0.52 |
| Congestive Heart Failure | 0.52 | Gastrointestinal |  | 0.58 | Pulmonology | 0.53 |
| Asthma | 0.52 | Oncologic |  | 0.59 | Primary Care | 0.53 |
| Appendicitis | 0.52 | Immunologic |  | 0.59 | Pediatrics | 0.53 |
| Autoimmune Disorders | 0.53 | Hepatic |  | 0.65 | Geriatrics | 0.54 |
| COPD | 0.53 | Infectious |  | 0.68 | Internal Medicine | 0.54 |
| Heart Failure | 0.54 | Renal |  | 0.68 | Surgery (General) | 0.54 |
| Endocarditis | 0.54 | Urinary |  | 0.78 | Hospitalist Medicine | 0.54 |
| Stroke/TIA | 0.54 |  |  |  | Anesthesiology | 0.54 |
| Chemotherapy | 0.54 |  |  |  | Critical Care | 0.56 |
| Atrial Fibrillation | 0.54 |  |  |  | Cardiology | 0.56 |
| Hypertension | 0.55 |  |  |  | Emergency Medicine | 0.56 |
| Respiratory Distress | 0.55 |  |  |  | Pain Management | 0.56 |
| COVID-19 | 0.56 |  |  |  | Neurology | 0.56 |
| Arrhythmia | 0.57 |  |  |  | Surgery (Cardiothoracic) | 0.57 |
| Dementia | 0.57 |  |  |  | Pediatric Subspecialty | 0.57 |
| Coronary Artery Disease | 0.57 |  |  |  | Radiology | 0.57 |
| Pneumonia | 0.58 |  |  |  | Critical Care (Neurologic) | 0.57 |
| Hepatitis | 0.58 |  |  |  | Critical Care (Pediatric) | 0.58 |
| GI Bleeding | 0.58 |  |  |  | Gastroenterology | 0.59 |
| Cirrhosis | 0.59 |  |  |  | Hematology and Oncology | 0.59 |
| Hematologic Malignancy | 0.59 |  |  |  | Palliative Care/Hospice | 0.59 |
| Bleeding/Hemorrhage | 0.59 |  |  |  | Pharmacy | 0.59 |
| Cancer | 0.60 |  |  |  | Toxicology | 0.59 |
| Idiopathic Pulmonary Fibrosis | 0.60 |  |  |  | Hepatology | 0.60 |
| Myocardial Infarction | 0.61 |  |  |  | Rehabilitation Medicine | 0.63 |
| Acute Coronary Syndrome | 0.61 |  |  |  | Infectious Disease | 0.63 |
| Leukemia/Lymphoma | 0.62 |  |  |  | Surgery (Trauma) | 0.63 |
| Alcoholism | 0.62 |  |  |  | Nephrology | 0.67 |
| Anemia | 0.64 |  |  |  | Urology | 0.83 |
| Trauma | 0.64 |  |  |  |  |  |
| Renal Failure | 0.66 |  |  |  |  |  |
| Drug/Alcohol Use | 0.67 |  |  |  |  |  |
| Hepatocellular Carcinoma | 0.71 |  |  |  |  |  |
| HIV/AIDS | 0.72 |  |  |  |  |  |

**Table A6 | CDIs that involve followup.**

| **Direct** | **Hybrid** | **Indirect** |
| --- | --- | --- |
| ABIC Score for Alcoholic Hepatitis | ARISCAT Score for Postoperative Pulmonary Complications | 4-Level Pulmonary Embolism Clinical Probability Score (4PEPS) |
| AWOL Score for Delirium | Aortic Dissection Detection Risk Score (ADD-RS) | ACTION ICU Score for Intensive Care in NSTEMI |
| Acute Gout Diagnosis Rule | CHADS-VASc Score for Atrial Fibrillation Stroke Risk | ALBI (Albumin-Bilirubin) Grade for Hepatocellular Carcinoma (HCC) |
| Altitude-Adjusted PERC Rule | Canadian Transient Ischemic Attack (TIA) Score | ATLAS Score for Clostridium Difficile Infection |
| Asthma Predictive Index (API) | Charlson Comorbidity Index (CCI) | ATRIA Bleeding Risk Score |
| BODE Index for COPD Survival | Dual Antiplatelet Therapy (DAPT) Score | ATRIA Stroke Risk Score |
| Binet Staging System for Chronic Lymphocytic Leukemia (CLL) | Emergency Department Assessment of Chest Pain Score (EDACS) | Alberta Stroke Program Early CT Score (ASPECTS) |
| CATCH (Canadian Assessment of Tomography for Childhood Head injury) Rule | HAS-BLED Score for Major Bleeding Risk | Asymptomatic Myeloma Prognosis |
| Canadian C-Spine Rule | HEART Pathway for Early Discharge in Acute Chest Pain | Blast Lung Injury Severity Score |
| Canadian CT Head Injury/Trauma Rule | HEART Score for Major Cardiac Events | Brain Metastasis Velocity (BMV) Model |
| Canadian Syncope Risk Score | HIT Expert Probability (HEP) Score for Heparin-Induced Thrombocytopenia | CHADS<sub>2</sub> Score for Atrial Fibrillation Stroke Risk |
| Columbia Suicide Severity Rating Scale (C-SSRS Screener) | Indian Takayasu Clinical Activity Score (ITAS2010) | CKD Prediction in HIV+ Patients |
| D'Amico Risk Classification for Prostate Cancer | Modified Hoehn and Yahr Scale for Parkinson’s Disease | CLIF-C ACLF (Acute-on-Chronic Liver Failure) |
| DIPSS (Dynamic International Prognostic Scoring System) for Myelofibrosis | PECARN Rule for Low Risk Febrile Infants 29-60 Days Old | COVID Home Safely Now (CHOSEN) Risk Score for COVID-19 |
| DRAGON Score for Post-TPA Stroke Outcome | Simplified PESI (Pulmonary Embolism Severity Index) | COVID-19 Inpatient Risk Calculator (CIRC) |
| Danger Assessment Tool for Domestic Abuse | Vancouver Chest Pain Rule | Clinical Disease Activity Index (CDAI) for Rheumatoid Arthritis |
| Disease Activity Score-28 for Rheumatoid Arthritis with CRP (DAS28-CRP) | Wells' Criteria for Pulmonary Embolism | DASH Prediction Score for Recurrent VTE |
| Disease Activity Score-28 for Rheumatoid Arthritis with ESR (DAS28-ESR) |  | DIRE Score for Opioid Treatment |
| Disease Steps for Multiple Sclerosis |  | Duval/CIBMTR Score for Acute Myelogenous Leukemia (AML) Survival |
| Duke Treadmill Score |  | EUTOS Score for Chronic Myelogenous Leukemia (CML) |
| EGSYS (Evaluation of Guidelines in SYncope Study) Score for Syncope |  | European System for Cardiac Operative Risk Evaluation (EuroSCORE) II |
| Endotracheal Tube (ETT) Depth and Tidal Volume Calculator |  | FINDRISC (Finnish Diabetes Risk Score) |
| Fong Clinical Risk Score for Colorectal Cancer Recurrence |  | Fibrosis-4 (FIB-4) Index for Liver Fibrosis |
| Functional Outcome in Patients With Primary Intracerebral Hemorrhage (FUNC) Score |  | Follicular Lymphoma International Prognostic Index (FLIPI) |
| Geneva Score (Revised) for Pulmonary Embolism |  | For Patients: VACO Index COVID-19 Mortality Risk |
| Groupe d'Etude des Lymphomes Folliculaires (GELF) Criteria |  | Fuhrman Nuclear Grade for Clear Cell Renal Carcinoma |
| HERDOO2 Rule for Discontinuing Anticoagulation in Unprovoked VTE |  | GALAD Model for Hepatocellular Carcinoma (HCC) |
| Hestia Criteria for Outpatient Pulmonary Embolism Treatment |  | GAP Index for Idiopathic Pulmonary Fibrosis (IPF) Mortality |
| LACE Index for Readmission |  | GRACE ACS Risk and Mortality Calculator |
| Lille Model for Alcoholic Hepatitis |  | Gail Model for Breast Cancer Risk |
| Liver Decompensation Risk after Hepatectomy for Hepatocellular Carcinoma (HCC) |  | Glasgow Prognostic Score (GPS) for Cancer Outcomes |
| Marburg Heart Score (MHS) |  | Gupta Postoperative Respiratory Failure Risk |
| Modified Asthma Predictive Index (mAPI) |  | HAT (Hemorrhage After Thrombolysis) Score for Predicting Post-tPA Hemorrhage |
| NEXUS Chest Decision Instrument for Blunt Chest Trauma |  | HEMORR<sub>2</sub>HAGES Score for Major Bleeding Risk |
| Ocular Hypertension Treatment Study (OHTS) Calculator |  | HINTS for Stroke in Acute Vestibular Syndrome |
| Ottawa Subarachnoid Hemorrhage (SAH) Rule for Headache Evaluation |  | HOSPITAL Score for Readmissions |
| PECARN Pediatric Head Injury/Trauma Algorithm |  | HScore for Reactive Hemophagocytic Syndrome |
| Padua Prediction Score for Risk of VTE |  | Hour-Specific Risk for Neonatal Hyperbilirubinemia |
| Palchak (UC Davis) Rule for Pediatric Head Trauma |  | Hypoglycemia Risk Score |
| Pediatric Asthma Severity Score (PASS) for Asthma Exacerbation Severity |  | IMDC (International Metastatic RCC Database Consortium) Risk Model for Metastatic Renal Cell Carcinoma |
| Pediatric NEXUS II Head CT Decision Instrument for Blunt Trauma |  | IMPACT Score for Outcomes in Head Injury |
| Prevention and Incidence of Asthma and Mite Allergy (PIAMA) Risk Score |  | IMPROVE RAM (International Medical Prevention Registry on Venous Thromboembolism Risk Assessment Model) |
| Psoriasis Area and Severity Index (PASI) |  | IMPROVE Risk Score for Venous Thromboembolism (VTE) |
| Revised Natural History Model for Primary Sclerosing Cholangitis |  | IMPROVEDD Risk Score for Venous Thromboembolism (VTE) |
| STONE Nephrolithometry Score for Renal Calculi |  | Infective Endocarditis (IE) Mortality Risk Score |
| Simple Disease Activity Index (SDAI) for Rheumatoid Arthritis |  | International Prognostic Index for Chronic Lymphocytic Leukemia (CLL-IPI) |
| Steinhart Model for Acute Heart Failure (AHF) in Undifferentiated Dyspnea |  | Khorana Risk Score for Venous Thromboembolism in Cancer Patients |
| Systemic Lupus Erythematosus Disease Activity Index 2000 (SLEDAI-2K) |  | Kidney Failure Risk Calculator |
| Troponin-only Manchester Acute Coronary Syndromes (T-MACS) Decision Aid |  | Killip Classification for Heart Failure |
| Truelove and Witts Severity Index for Ulcerative Colitis |  | Kruis Score for Diagnosis of Irritable Bowel Syndrome (IBS) |
| Urticaria Activity Score (UAS) |  | LENT Prognostic Score for Malignant Pleural Effusion |
| WPSS (WHO classification-based Prognostic Scoring System) for Myelodysplastic Syndrome |  | Leiden Clinical Prediction Rule for Undifferentiated Arthritis |
| YEARS Algorithm for Pulmonary Embolism (PE) |  | MAGGIC Risk Calculator for Heart Failure |
|  |  | MELD Na (UNOS/OPTN) |
|  |  | MELD Score (Original, Pre-2016, Model for End-Stage Liver Disease) |
|  |  | Mantle Cell Lymphoma International Prognostic Index (MIPI) |
|  |  | Mekhail Extension of the Motzer Score |
|  |  | Memorial Sloan-Kettering Cancer Center (MSKCC/Motzer) Score for Metastatic Renal Cell Carcinoma (RCC) |
|  |  | Metroticket Calculator for Hepatocellular Carcinoma (HCC) Survival |
|  |  | Michigan Risk Score for PICC-Related Thrombosis |
|  |  | Mirels’ Criteria for Prophylactic Fixation |
|  |  | Modified Glasgow Prognostic Score (mGPS) for Cancer Outcomes |
|  |  | Modified NIH Stroke Scale/Score (mNIHSS) |
|  |  | Montreal Classification for Inflammatory Bowel Disease (IBD) |
|  |  | Morphine Milligram Equivalents (MME) Calculator |
|  |  | Mumtaz Score for Readmission in Cirrhosis |
|  |  | NEXUS Chest CT Decision Instrument for CT Imaging |
|  |  | Nutritional Risk Index (NRI) |
|  |  | ORBIT Bleeding Risk Score for Atrial Fibrillation |
|  |  | Opioid Risk Tool (ORT) for Narcotic Abuse |
|  |  | Ottawa COPD Risk Scale |
|  |  | Ottawa Heart Failure Risk Scale (OHFRS) |
|  |  | PEDIS Score for Diabetic Foot Ulcers |
|  |  | PLASMIC Score for TTP |
|  |  | POMPE-C Tool for Pulmonary Embolism Mortality |
|  |  | POSSUM for Operative Morbidity and Mortality Risk |
|  |  | PREVAIL Model for Prostate Cancer Survival |
|  |  | PRIEST COVID-19 Clinical Severity Score |
|  |  | PSA Doubling Time (PSADT) Calculator |
|  |  | Patient Activity Scale (PAS) for RA |
|  |  | Patient Activity Scale II (PAS II) for RA |
|  |  | Pittsburgh Response to Endovascular therapy (PRE) Score |
|  |  | Preoperative Mortality Predictor (PMP) Score |
|  |  | Prognostic Index for Cancer Outcomes |
|  |  | Pulmonary Embolism Severity Index (PESI) |
|  |  | RCVS<sub>2</sub> Score for Reversible Cerebral Vasoconstriction Syndrome |
|  |  | REACH-B Score for Hepatocellular Carcinoma (HCC) |
|  |  | READMITS Score for Readmissions in Acute MI |
|  |  | REVEAL Registry Risk Score 2.0 for Pulmonary Arterial Hypertension (PAH) |
|  |  | ROSE (Risk Stratification of Syncope in the Emergency Department) Rule |
|  |  | Rai Staging System for Chronic Lymphocytic Leukemia (CLL) |
|  |  | Recurrent Instability of the Patella (RIP) Score |
|  |  | RegiSCAR Score for Drug Reaction with Eosinophilia and Systemic Symptoms (DRESS) |
|  |  | Revised International Prognostic Scoring System (IPSS-R) for Myelodysplastic Syndrome (MDS) |
|  |  | Revised Multiple Myeloma International Staging System (R-ISS) |
|  |  | Reynolds Risk Score for Cardiovascular Risk in Women |
|  |  | Risk of Paradoxical Embolism (RoPE) Score |
|  |  | Salivary Gland Cancer Model for Survival +/- Postoperative Radiotherapy (PORT) |
|  |  | Simon Broome Diagnostic Criteria for Familial Hypercholesterolemia (FH) |
|  |  | Sokal Index for Chronic Myelogenous Leukemia (CML) |
|  |  | THRIVE Score for Stroke Outcome |
|  |  | Therapy-Disability-Neurology (TDN) Grade |
|  |  | UCLA Integrated Staging System (UISS) for Renal Cell Carcinoma (RCC) |
|  |  | UCSF-CAPRA Score for Prostate Cancer Risk |
|  |  | United Kingdom Model for End-Stage Liver Disease (UKELD) |
|  |  | Veterans Aging Cohort Study (VACS) 1.0 Index |
|  |  | Veterans Health Administration COVID-19 (VACO) Index for COVID-19 Mortality |

**Table A7 | CDIs without retrievable primary literature**

| **CDI Name** | **Description** |
| --- | --- |
| Bicarbonate Deficit | Calculates total body bicarb deficit. |
| LDL Calculated | From Total Chol, HDL and Trigs. |
| Pregnancy Due Dates Calculator | From LMP, EGA, or date of conception. |
| Steroid Conversion Calculator | Steroid dosing equivalencies. |
| Gleason Score for Prostate Cancer | Prognoses prostate cancer. |
| Immunization Schedule Calculator | Immunizations due based on age. |
| Schwab and England Activities of Daily Living (ADL) Scale | Parkinson's disease disability. |
| New York Heart Association (NYHA) Functional Classification for Heart Failure | Stratifies severity of heart failure. |
| DSM-5 Criteria for Binge Eating Disorder | Diagnosis of BED. |
| Pack Years Calculator | Quantify smoking history. |
| DSM-5 Criteria for Major Depressive Disorder | Diagnosis of MDD. |
| DSM-5 Criteria for Bipolar Disorder | Diagnosis of BPD. |
| DSM-5 Criteria for Posttraumatic Stress Disorder | Diagnosis of PTSD. |
| Utah COVID-19 Risk Score | Oral antiviral recommendation. |
| Indications for Paxlovid | Paxlovid indication. |
| Revised Urinary Incontinence Scale (RUIS) | Assesses for incontinence. |
| Urine Output and Fluid Balance | Urine output over 24 hrs. |
| Atropine Dosing for Cholinesterase Inhibitor Toxicity | Cholinesterase inhibitor reversal. |
| Karnofsky Performance Status Scale | Chemotherapy tolerance. |
| Estimated Ethanol (and Toxic Alcohol) Serum Concentration Based on Ingestion | Predicts ethanol concentration based on ingestion of alcohol. |
| Dutch Criteria for Familial Hypercholesterolemia (FH) | Diagnoses familial hypercholesterolemia. |
| Global Initiative for Obstructive Lung Disease (GOLD) Criteria for COPD | COPD staging and recommendations. |
| High-dose Insulin Euglycemia Therapy (HIET) | Calcium-channel blocker, beta blocker reversal. |
| Fomepizole Dosing | Methanol and ethylene glycol reversal. |
| Sickle Cell RBC Exchange Volume | Donor RBC volume for exchange. |
| Cryoprecipitate Dosing for Fibrinogen Replacement | Cryoprecipitate dosing. |
| Maternal-Fetal Hemorrhage Rh(D) Immune Globulin Dosage | RhIG for HDFN. |
| RBC Units to Screen for Compatibility | Donors to antigen type. |
| Donor Lymphocyte Infusion (DLI) Volume | Estimates DLI dosage. |
| Modified Finnegan Neonatal Abstinence Score (NAS) | Opioid withdrawal in newborns. |
| Visual Acuity Testing (Snellen Chart) | Visual acuity. |
| Color Vision Screening (Ishihara Test) | Color blindness. |
| Benzodiazepine Conversion Calculator | Benzodiazepine conversion. |
| Cardiac Output (Fick’s Formula) | Calculates CO, CI, and SV. |
| Villalta Score for Post-thrombotic Syndrome (PTS) | Post-thrombotic syndrome severity. |
| Local Anesthetic Dosing Calculator | Local anesthetic toxicity prevention. |
| Brescia-COVID Respiratory Severity Scale (BCRSS)/Algorithm | COVID-19 pathway/approach from Italy. |
| ACEP ED COVID-19 Management Tool | ED management tool for COVID-19. |
